# Non applicability of validated predictive models for intensive care admission and death of COVID-19 patients in a secondary care hospital in Belgium

**DOI:** 10.1101/2020.11.06.20205799

**Authors:** Nicolas Parisi, Aurore Janier-Dubry, Ester Ponzetto, Charalambos Pavlopoulos, Gaetan Bakalli, Roberto Molinari, Stéphane Guerrier, Nabil Mili

## Abstract

**Objective:** To set up simple and reliable predictive scores for intensive care admissions and deaths in COVID-19 patients. These scores adhere to the TRIPOD (transparent reporting of a multivariable prediction model for individual prognosis or diagnosis) reporting guidelines.

**Design:** Monocentric retrospective cohort study run from early March to end of May in Clinique Saint-Pierre Ottignies, a secondary care hospital located in Ottignies-Louvain-la-Neuve, Belgium. The outcomes of the study are (i) admission in the Intensive Care Unit and (ii) death.

**Data sources:** All patients admitted in the Emergency Department with a positive RT-PCR SARS-CoV-2 test were included in the study. Routine clinical and laboratory data were collected at their admission and during their stay. Chest X-Rays and CT-Scans were performed and analyzed by a senior radiologist.

**Methods:** A recently published predictive score conducted on a large scale was used as a benchmark value (Liang score)^1^. Logistic regressions were used to develop predictive scores for (i) admission to ICU among emergency ward patients; (ii) death among ICU patients on 40 clinical variables. These models were based on medical intuition and simple model selection tools. Their predictive capabilities were then compared to Liang score.

**Results:** Our results suggest that Liang score may not provide reliable guidance for ICU admission and death. Moreover, the performance of this approach is clearly outperformed by models based on simple markers. For example, a logistic regression considering only the LDH yields to similar sensitivity and greater specificity. Finally, all models considered in this study lead to levels of specificity under or equal to 50%.

**Conclusions:** In our experience, the results of a predictive score based on a large-scale Chinese study cannot be applied in the Belgian population. However, in our small cohort it appears that LDH above 579 UI/L and venous lactate above 3.02 mmol/l may be considered as good predictive biological factors for ICU admission. With regards to death risk, NLR above 22.1, tobacco abuse status and <u>80 % of</u> respiratory impairment appears to be relevant predictive factors. A predictive score for admission to ICU or death is desperately needed in secondary hospitals. Optimal allocation of resources guided by evidence-based indicators will best guide patients at time of admission and avoid futile treatments in intensive care units.

## Introduction

The novel coronavirus disease 2019 (COVID-19) presents an important and urgent threat to global health. Since the outbreak in early December 2019 in the Hubei province of the People’s Republic of China, the number of patients confirmed to have the disease has exceeded 31,717,955 in more than 188 countries (as of 23 September 2020), and the number of people infected is probably much higher^2^. More than 973,014 people have died from COVID-19 infection (as of 23 September 2020)^2^. Despite public health responses aimed at containing the disease and delaying the spread, several countries including Belgium have been confronted with a critical care crisis. Outbreaks lead to important increases in the demand for hospital beds and shortage of medical equipment. The adequacy of resources to treat infected cases is therefore a growing public health concern.

The spread of COVID-19 has been heterogeneous, resulting in some regions having sporadic transmission and relatively few hospitalized patients with COVID-19 and others having community transmission that has led to overwhelming numbers of severe cases. The latter is the case for Belgium which has one of the highest incidence and death rates in the world (9,955 deaths, corresponding to 858 deaths per million inhabitants (up to 23 September 2020)^2^. South of Brussels, in a region called Brabant Wallon in Belgium, where Clinique Saint-Pierre Ottignies is located, healthcare delivery has been compromised by critical resource constraints in diagnostic testing, hospital beds, ventilators, and healthcare workers. Optimal allocation of resources has led us to build easily implemented clinical indicators in order to best guide patients at the time of admission and avoid futile treatments in intensive care units.

Time is a huge constraint in the management of COVID-19 patients during the pandemic. We excluded from our study all biological variables that were not available within a few hours after blood sampling. In doing so, we are in line with the reality of most non-tertiary care hospitals in Europe.

### Radiology

Thoracic imaging with chest radiography (CXR) and thoracic computed tomography (TCT) are key tools for pulmonary disease diagnosis and management, but their role in the management of COVID-19 has been sparsely considered within the multivariable context of the severity of respiratory disease, pre-test probability, risk factors for disease progression, and critical resource constraints. In a recent consensus article^3^, a multidisciplinary panel comprised principally of radiologists and pulmonologists from 10 countries with experience managing COVID-19 patients across a spectrum of healthcare environments has recently evaluated the utility of imaging within three scenarios representing varying risk factors, community conditions, and resource constraints. The main recommendations of this panel of experts are:

- Imaging is not routinely indicated as a screening test for COVID-19 in asymptomatic Individuals;
- Imaging is not indicated for patients with mild features of COVID-19 unless they are at risk for disease progression;
- Imaging is indicated for patients with moderate to severe features of COVID-19 regardless of COVID-19 test results;
- Imaging is indicated for patients with COVID-19 and evidence of worsening respiratory status;
- In a resource-constrained environment where access to CT is limited, CXR may be preferred for patients with COVID-19 unless features of respiratory worsening warrant the use of CT.

These recommendations, quite sensible at first sight, are not confirmed by any study and are challenged by our clinical experience. A recent review of the radiological manifestations of COVID-19 gives no clear-cut conclusions^4^ and the association between radiological findings and outcome remain elusive. We consequently included two radiological variables within the features of interest and checked their statistical significance.

### Tripod statements

A recent publication^5^ provides a review and critical appraisal of published and preprint reports of prediction models for diagnosing patients with suspected infection, for prognosis of COVID-19 patients, and for detecting people in the general population at risk of being admitted to hospital for COVID-19 pneumonia. In this review, 31 prediction models were included. The conclusions of the authors are clear-cut: all studies are rated at high risk of bias, mostly because of non-representative selection of control patients, exclusion of patients who had not experienced the event of interest by the end of the study, and high risk of model overfitting. Reporting quality varied substantially between studies. Most reports did not include a description of the study population or intended use of the models, and calibration of predictions was rarely assessed. Authors recommended that further studies should adhere to the TRIPOD (transparent reporting of a multivariable prediction model for individual prognosis or diagnosis) reporting guideline^6^.

### Clinical score in the literature

Among the clinical scores available in the literature, one of them drew our attention^1^. Having been recently published in JAMA *Internal Medicine*, it addresses the same issues as our study, but was conducted on a much larger scale. According to the authors, the development cohort included 1,590 patients. The mean (SD) age of patients in this cohort was 48.9 (15.7) years; 904 (57.3%) were men. The validation cohort included 710 patients with a mean (SD) age of 48.2 (15.2) years, and 382 (53.8%) were men and 172 (24.2%). From 72 potential predictors, 10 variables were independent predictive factors and were included in the risk score: chest radiographic abnormality (OR, 3.39; 95%CI, 2.14-5.38), age (OR, 1.03; 95%CI, 1.01-1.05), hemoptysis (OR, 4.53; 95%CI, 1.36-15.15), dyspnea (OR, 1.88; 95%CI, 1.18-3.01), unconsciousness (OR, 4.71; 95%CI, 1.39-15.98), number of comorbidities (OR, 1.60; 95%CI, 1.27-2.00), cancer history (OR, 4.07; 95%CI, 1.23-13.43), neutrophil-to-lymphocyte ratio (OR, 1.06; 95%CI, 1.02-1.10), lactate dehydrogenase (OR, 1.002; 95%CI, 1.001-1.004) and direct bilirubin (OR, 1.15; 95%CI, 1.06-1.24). The Area Under the ROC Curve (AUC) in the development cohort was 0.88 (95%CI, 0.85-0.91) and the AUC in the validation cohort was 0.88 (95%CI, 0.84-0.93). The score has been translated into an online risk calculator that is freely available (http://118.126.104.170/). The fact that the clinical score was validated on an independent large population gives some credit to its applicability on a large scale. We decided then to include this score in our study and, for each of our COVID-19 patients, we calculated the clinical risk score at time d_0_ (day of admission to the emergency ward) using the freely available calculator. We refer to this clinical risk score as Liang score. Liang score prediction accuracy is used as a benchmark of our own search of a predictive model. However, in contrast to Liang approach, we have distinguished two outcomes that cannot be confounded: admission to ICU and death.

## Research questions

1. Is it possible to predict admission of COVID-19 patients to ICU with routine and quickly available clinical, biological and radiological variables? What is the prediction error of the selected model(s)?
2. Is it possible to predict death among COVID-19 patients with routine and quickly available clinical, biological and radiological variables, in order to avoid unnecessary treatment and waste of precious resources? What is its prediction error?

## Methods

We conducted a retrospective cohort study of 66 patients with known COVID-19 disease, from March 10^th^ to May 16^th^ 2020 in Clinique Saint-Pierre Ottignies in Belgium. Twenty-two patients admitted in Intensive Care Unit and 44 in Medicine Department. 63 patients were positive for a nasopharyngeal RT-PCR SARS-CoV-2 test. One patient included in the study was negative for RT-PCR SARS-CoV-2 but had an IgA and IgG ELISA-test positive for SARS-CoV-2 and ground-glass opacities on the chest X-ray.

All patients were followed from their admission at the emergency ward until they get out of the hospital or until their death. No patient was excluded from the cohort. The follow-up ended when patients leaved the hospital or died. The starting date of accrual and the end-date of accrual were reported for each patient.

This hospital is a 425-bed regional general hospital with a capacity of 15 intensive care beds, which was increased to 25 beds during the pandemic. Clinique Saint-Pierre Ottignies has a mission of para-university training of junior medical specialists. It covers a catchment area of circa 400,000 patients, in the region of Brabant Wallon (Wallonia, Belgium).

### Criteria of admission to the ICU

Twenty-two patients with COVID-19 were admitted to the ICU. For 19 patients the reason was a respiratory failure defined as (i) ambient oxygen saturation (SpO_2_) < 88% with nasal cannula oxygen therapy > 5l/min; (ii) PaO_2_ < 50 mmHg and/or a ratio PaO_2_/FiO_2_ < 150; (iii) respiratory rate > 40/min. For one patient, the reason was a post-traumatic cerebral hemorrhage, associated with altered neurological status (defined as Glasgow coma scale < 8/15) requiring mechanical ventilation in order to protect the airway. For another one, the cause was the postoperative management of an empyema drainage developed in a context of bacterial pneumonia complicating a SARS-CoV-2 virus infection. A third patient presented a status epilepticus in the context of probable alcohol withdrawal.

### Criteria of non-admission to the ICU

Clinique Saint-Pierre Ottignies has long-proven guide-lines for admission to ICU. Given the nature of the health emergency, the principle of distributive justice^7^ was applied and each patient admitted to the emergency department was immediately classified as “eligible for intensive care” or “not eligible for intensive care”, taking into account his or her previous history and quality of life. The criteria for ineligibility were: (i) presence of a prior incurable disease; (ii) limitation of functional autonomy; and (iii) advanced dementia. Two patients had criteria for intensive care hospitalization upon admission to the emergency room, the other patients were first hospitalized in a non-intensive care unit.

The patients admitted to intensive care all had the clinical criteria mentioned above, with a pre-established maximalist therapeutic plan. Other patients with the same clinical criteria but with a care plan with therapeutic limitations were not admitted. These therapeutic projects were discussed collegially between medical specialties issued from emergency department, internal medicine and critical unit.

The advantage of a simple predictive score upon admission could be useful in decision-making regarding a therapeutic plan.

### Assessment of lung injury

A senior radiologist analyzed Thoracic Computer Tomographies (TCT) according to two criteria: percentage of lung injury (continuous variable from 0 to 100%) and density of lung injury (factor variable with 3 grades: 1 = light density; 2 = moderate density; 3 = high density).

### Data collection

Data collection tried to adhere as tightly as possible to the TRIPOD Adherence extraction form (https://www.tripod-statement.org/wp-content/uploads/2020/03/TRIPOD-Adherence-assessment-form_V-2018_12.pdf). At their admission, patients were questioned about their usual medication and their health condition. The body mass index was computed. Collected variables are the following: age, gender, ethnic group, weight, body mass index, number of days with symptoms before hospitalization, asthenia, pyrexia, dyspnea, chest pain, aspecific digestive symptoms, anosmia, ageusia, confusion, Travel or contact < one month, cigarette consumption (Y or N), hypertension, diabetes, mental status (depression), angiotensin-converting-enzyme inhibitors, angiotensin II receptor antagonists, non-steroidal anti-inflammatory drugs, immunosuppressive drugs, SpO_2_ (%), TCT % of lung injury, TCT density of lung injury, blood type, white blood cells, neutrophils, lymphocytes, blood platelets, fibrinogen, ferritin, triglycerides, LDH, troponin, CRP, neutrophil-to-lymphocyte ratio, lymphocyte-to-CRP ratio, bilirubin and lactates. The dates of admission to ICU and death were recorded.

In the ICU, the use of chloroquine, hydroxychloroquine, azithromycin, clarithromycin, remdesevir and antibacterial antibiotics (piperacill-tazobactam, meropenem, ciprofloxacin, ceftazidime, amoxicillin clavulanate) was recorded on a daily basis.

As mentioned previously, the clinical risk score developed by Liang et al^1^ was computed retrospectively for all COVID-19 patients at their admission to the emergency ward. This clinical risk score was compared to our own models.

### Statistical analysis

The number of collected variables being 40, the number of possible predictive models^8^ is extremely large. Consequently, testing all of these models is not computationally feasible and a preselection of potential predictive models was performed using a recently developed method^9^. Among these models, we decided to consider four models that were in line with medical practice and presented suitable prediction accuracy for ICU admission. These models are logistic regressions based on the following variables: (i) LDH, (ii) LDH + sex, (iii) LDH + sex + venous lactate, and (iv) respiratory impairment score (a combination of percentage of lung injury, density of lung injury, and ambient oxygen saturation). Similarly, three models presented were considered death: (i) Neutrophil Lymphocyte Ratio + Tobacco; (ii) Liang score + Neutrophil Lymphocyte Ratio + Tobacco; and (iii) respiratory impairment score.

As previously mentioned, logistic regressions were used in our study and this techniques allows to addressed to our research questions based on binary variables. All analysis were performed using the R software version 4.0.1 (https://cran.r-project.org/). This regression technique is a widely used statistical tool that allows for multivariate analysis and modeling of a binary dependent variable. The multivariate analysis estimates coefficients (for example, log odds or hazard ratios) for each predictor included in the final model and adjusts them with respect to the other predictors in the model. The coefficients quantify the contribution of each predictor to the outcome risk estimation^10^. The caveats to consider when assessing the results of a logistic regression analysis are well explained in Tolles *et al*.^11^. Theoretically, every variable collected in the study could be a candidate predictor. However, to reduce the risk of false positive findings and improve model performance, the Events Per Variable (EPV) must be considered. A rule of thumb of 10 individuals per event is commonly applied^12^. However recent studies have shown that EPV does not have a strong relation with metrics of predictive performance, and is not an appropriate criterion for binary prediction model development studies^13^. According to Vittinghof *et al*.^14^, predictive performance problems are fairly frequent with 2–4 EPV, uncommon with 5–9 EPV, and still observed with 10–16 EPV. The rule of thumb of 10 individuals per event can then be relaxed. In our study, EPV ranges from 7 to 20. The binomial family logit function and the maximum likelihood approach were used to compute the regression coefficients. The coefficients are then equivalent to the relative risk of the outcome in the exposed group (COVID-19 patients).

## Results

Admission to ICU and death are the relevant end-points of our study, and they have been handled separately. Pathophysiology of COVID-19 is complex^15^ and the process leading to death merges intrinsic factors (endothelial dysfunction^16^, thrombotic complications^17^, respiratory distress syndrome^18^, renal failure^19^, cardiovascular collapse^20^, etc.) and extrinsic factors to the disease (patient self-inflicted lung injury^21^, central nervous system impairment due to long-term sedation^22^, etc.). Moreover, admission to ICU and death are outcomes that are deeply different in nature. This point will be discussed further on. For both outcomes, Liang score is the benchmark to which any predictive score will be compared to.

### Admission to Intensive Care Unit

Predictive power of Liang score on the Belgian cohort was computed and used as a reference standard for our selected models. Results are shown in Table 1. If the sensitivity of Liang score value is high, its specificity is very low (0.10). Its regression coefficient is close to zero and above the significance threshold of 5% (cf. Table 2). Liang score predicts 2 admissions to ICU out of 20 patients and cannot reliably be used as a clinical tool in the emergency ward. In the wake of this disappointing result, we computed our own predictive scores, with a minimum number of covariates in order to keep the EPV in the 7 to 10 range. Based on clinical experience and on recent recommandations^5^, Sex, LDH, Neutrophil to Lymphocyte Ratio (NLR), venous lactate, SpO_2_, and respiratory impairment were selected as potential predictive features. The respiratory impairment score is built out of three variables: percentage of lung injury (continuous variable from 0 to 100%), density of lung injury (factor variable with 3 grades: 1 = light density; 2 = moderate density; 3 = high density), and SpO_2_. Our predictive models are respectively: (i) LDH; (ii) LDH + Sex; (iii) LDH + Sex + Lactate; and (iv) Respiratory impairment. The estimated parameters for these models are presented in Tables 3 to 6.

**Table 1.** Predictive performances of competing models regarding admission to ICU.

|  | Liang et al. score | LDH | LDH + Sex | LDH + Sex +<br>Lactate | Respiratory<br>impairment |
| --- | --- | --- | --- | --- | --- |
| **Accuracy** | 0.67<br>(0.54-0.78) | 0.78<br>(0.66-0.87) | 0.77<br>(0.64-0.86) | 0.80<br>(0.68-0.89) | 0.77<br>(0.64-0.86) |
| **Sensitivity** | 0.93 | 0.95 | 0.89 | 0.93 | 0.95 |
| **Specificity** | 0.10 | 0.40 | 0.50 | 0.50 | 0.35 |
| **PPV** | 0.69 | 0.78 | 0.80 | 0.80 | 0.76 |
| **NPV** | 0.40 | 0.80 | 0.67 | 0.77 | 0.78 |
| **Prevalence** | 0.69 | 0.69 | 0.69 | 0.69 | 0.69 |
| **EPV** | 20 | 20 | 10 | 7 | 7 |

PPV: positive predictive value; NPV: negative predictive value; EPV: events per variable. Inside brackets : Accuracy 95% confidence intervals.

**Table 2.** Liang score logistic regression coefficients (admission to ICU)

|  | Estimate | Std. error | z value | Pr (>\|z\|) |
| --- | --- | --- | --- | --- |
| Intercept | -2.85 | 1.17 | -2.44 | 0.0145 * |
| Score | 0.015 | 0.008 | 1.85 | 0.06 |

p-value : '\*\*\*\*' 0.001 ; '\*\*\*' 0.01 ; '\*' 0.05.

**Table 3.**
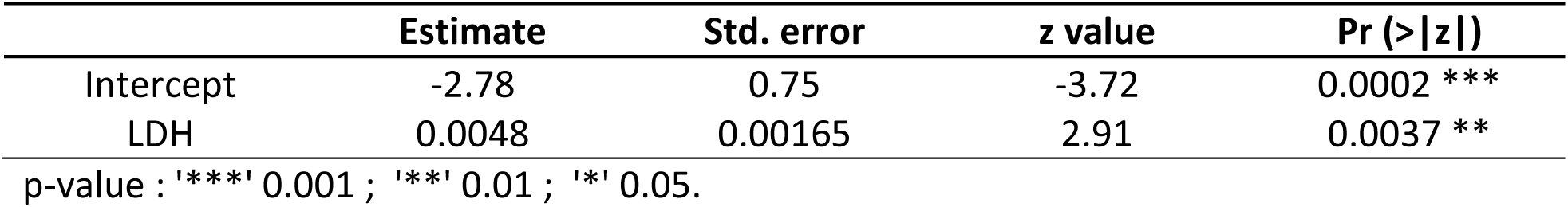
LDH score logistic regression coefficients (admission to ICU)

**Table 4.** (LDH + Sex) score logistic regression coefficients (admission to ICU)

|  | Estimate | Std. error | z value | Pr (>\|z\|) |
| --- | --- | --- | --- | --- |
| Intercept | -3.95 | 1.01 | -3.89 | 9.86e-05 *** |
| LDH | 0.0048 | 0.00172 | 2.78 | 0.0054 ** |
| Sex (M) | 1.71 | 0.754 | 2.26 | 0.0236 * |

p-value : '\*\*\*\*' 0.001 ; '\*\*\*' 0.01 ; '\*' 0.05.

**Table 5.** (LDH + Sex + venous lactate) score logistic regression coefficients (admission to ICU)

|  | Estimate | Std. error | z value | Pr (>\|z\|) |
| --- | --- | --- | --- | --- |
| Intercept | -6.02 | 1.55 | -3.89 | 0.0001 *** |
| LDH | 0.0055 | 0.0020 | 2.72 | 0.0065 ** |
| Sex (M) | 1.87 | 0.832 | 2.25 | 0.0241 * |
| Venous lactate | 0.86 | 0.34 | 2.55 | 0.011 * |

p-value : '\*\*\*\*' 0.001 ; '\*\*\*' 0.01 ; '\*' 0.05.

**Table 6.** Respiratory impairment score logistic regression coefficients (admission to ICU)

|  | Estimate | Std. error | z value | Pr (>\|z\|) |
| --- | --- | --- | --- | --- |
| Intercept | 7.64 | 5.64 | 1.35 | 0.175 |
| % LI | 0.022 | 0.016 | 1.373 | 0.17 |
| DLI | -0.172 | 0.41 | -0.421 | 0.67 |
| SpO<sub>2</sub> | -0.101 | 0.0595 | -1.699 | 0.89 |

% LI: percentage of lung injury; DLI: density of lung injury; SpO<sub>2</sub>: ambient oxygen saturation p-value: '\*\*\*\*' 0.001 ; '\*\*\*' 0.01 ; '\*' 0.05.

It appears that no score is quite satisfactory to predict admission to ICU. All scores that were tested are flawed by a lack of specificity and cannot be reliably used in the emergency ward. However, the absence of significant results does not mean that no valuable information can be extracted. Indeed, there are two kinds of useful information that can be found in these results.

First, a predictive score designed on a development cohort of 1,590 patients and validated over 710 patients^1^ in China appears to perform poorly on a European population with overall accuracy lower than 70% [95% CI: 0.5431-0.7841], and specificity around 10%. This surprising result requires additional clarification. At first sight, the main reason of Liang score poor performance on the Belgian cohort lies in the difference of end-points. Based on the American Thoracic Society guidelines for community-acquired pneumonia^23^, Liang *et al*. defined critical COVID-19 illness as a composite of admission to the intensive care unit, invasive ventilation, or death. They adopted this composite end-point for the reason that admission to ICU, invasive ventilation, and death are serious outcomes of COVID-19 that have been adopted in previous studies to assess the severity of other serious infectious diseases. However admission to ICU, invasive ventilation, and death are end-points whose nature is quite different. Admission to ICU and invasive ventilation depend on many extrinsic factors such as availability of intensive care beds, hospital care policy, and fair allocations of resources. These factors can vary widely over time. At the peak of the pandemic, overcrowding of ICU beds can divert patients to lighter care structures. On the other hand, death could be seen as a more “objective” end-point, less prone to health care policy fluctuations. The difference of end-points may then explain the lack of predictive power for Liang score. Another reason may lie in the genetics of population. This point deserves further research.

The second information is that respiratory impairment performs better than Liang score but can predict only 7 admissions to ICU out of 20. This finding confirms the fact that COVID-19 is much more than a respiratory distress syndrome^24^; it is essentially a multiple endothelial dysfunction^25^ with de novo angiogenesis and thrombosis^26^.

### Death

A series of predictive scores were tested along with that put forward in Liang *et al*. Their performances are displayed in Table 7. Surprisingly, the LDH does not appear to predict death reliably while it was the central variable for ICU admission. The only factor that seems to prodive reliable information. Indeed, a logistic regression based on this variable outperforms Liang score but its specificity remains low (24%). Two other models were considered: (i) NLR + Tobacco, (ii) NLR + Tobacco + Liang score, which provides similar predictive results. Logistic regression coefficients are displayed in Tables 8 to 11.

**Table 7.** Predictive performances of competing models regarding death.

|  | Liang score | NLR +<br>Tobacco | NLR + Tobacco +<br>Liang score | Respiratory<br>impairment |
| --- | --- | --- | --- | --- |
| **Accuracy** | 0.67<br>(0.54-0.78) | 0.69<br>(0.56-0.80) | 0.69<br>(0.56-0.80) | 0.77<br>(0.64-0.86) |
| **Sensitivity** | 0.93 | 0.91 | 0.89 | 0.96 |
| **Specificity** | 0.10 | 0.20 | 0.25 | 0.24 |
| **PPV** | 0.69 | 0.71 | 0.72 | 0.78 |
| **NPV** | 0.40 | 0.50 | 0.50 | 0.67 |
| **Prevalence** | 0.69 | 0.69 | 0.69 |  |
| **EPV** |  |  |  |  |

PPV: positive predictive value; NPV: negative predictive value; EPV: events per variable; NLR: Neutrophil-lymphocyte ratio. Inside brackets: Accuracy 95% confidence interval

**Table 8.** Liang score logistic regression coefficients (death)

|  | Estimate | Std. error | z value | Pr (>\|z\|) |
| --- | --- | --- | --- | --- |
| Intercept | -3.91 | 1.31 | -2.97 | 0.0029** |
| Score | 0.021 | 0.009 | 2.325 | 0.02* |

p-value : '\*\*\*\*' 0.001 ; '\*\*\*' 0.01 ; '\*' 0.05.

**Table 9.**
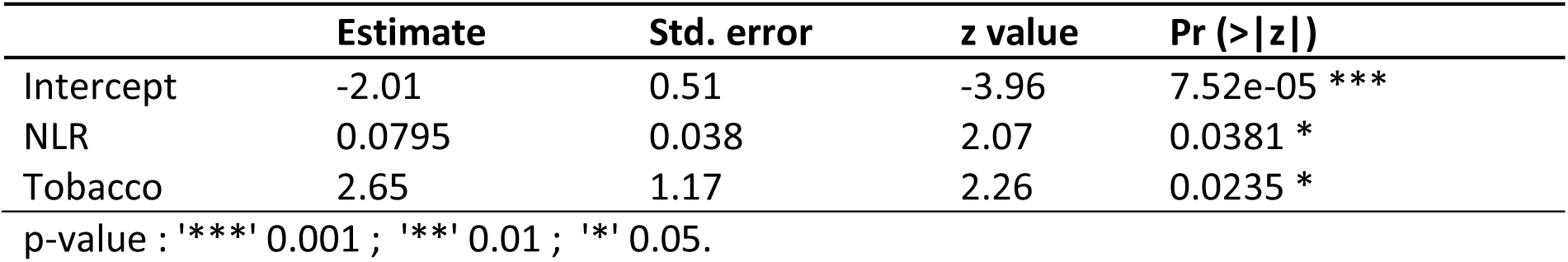
(NLR + Tobacco) score logistic regression coefficients (death)

|  | Estimate | Std. error | z value | Pr (>\|z\|) |
| --- | --- | --- | --- | --- |
| Intercept | -2.01 | 0.51 | -3.96 | 7.52e-05 *** |
| NLR | 0.0795 | 0.038 | 2.07 | 0.0381 * |
| Tobacco | 2.65 | 1.17 | 2.26 | 0.0235 * |

p-value : '\*\*\*\*' 0.001 ; '\*\*\*' 0.01 ; '\*' 0.05.

**Table 10.** (Liang + NLR + Tobacco) score logistic regression coefficients (death)

|  | Estimate | Std. error | z value | Pr (>\|z\|) |
| --- | --- | --- | --- | --- |
| Intercept | -3.94 | 1.40 | -2.82 | 0.0049** |
| Liang | 0.016 | 0.010 | 1.55 | 0.12 |
| NLR | 0.053 | 0.042 | 1.27 | 0.20 |
| Tobacco | 2.64 | 1.197 | 2.20 | 0.0275 * |

p-value : '\*\*\*\*' 0.001 ; '\*\*\*' 0.01 ; '\*' 0.05.

**Table 11.**
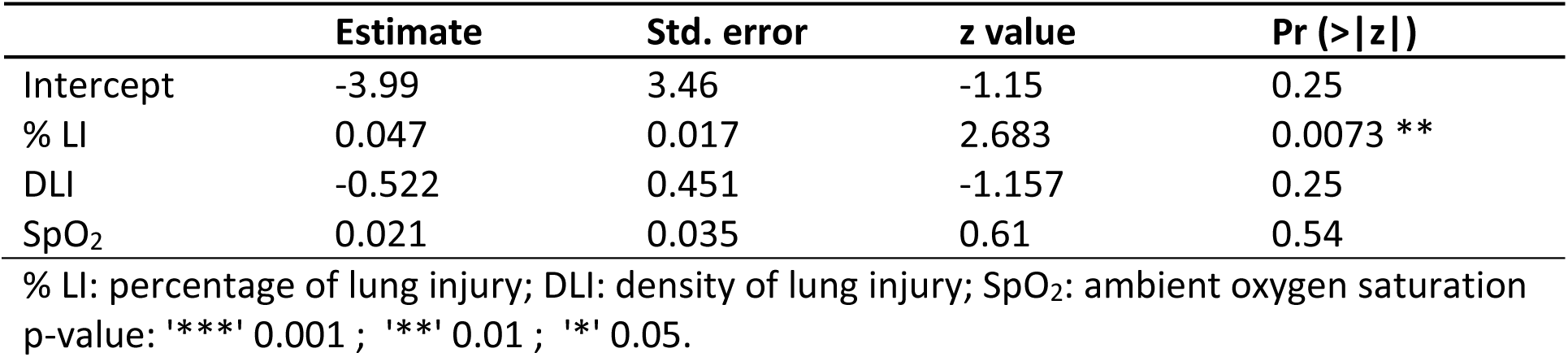
Respiratory impairment score logistic regression coefficients (death)

**Table 12.**
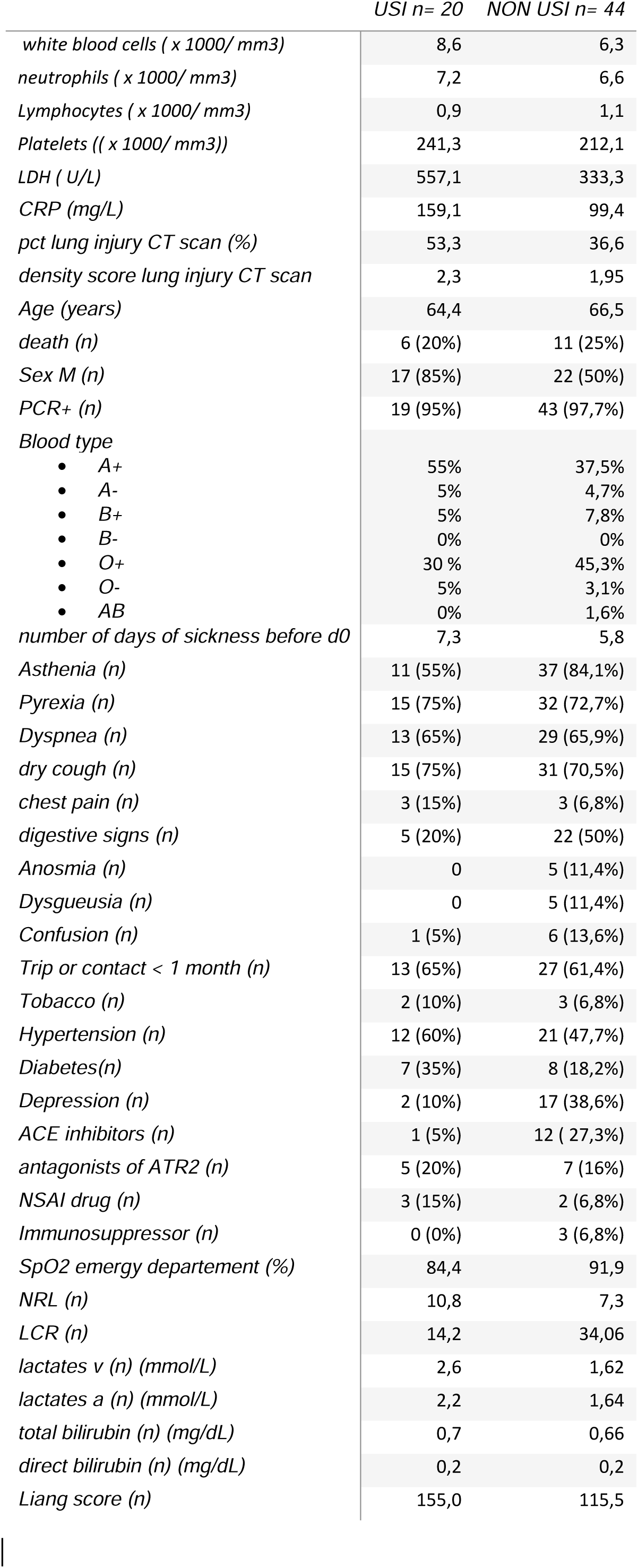
Characteristics of the patients.

#### The cut-off of percentage of lung injury that predicts death is 80 %

Similar conclusions can be drawn as those made for ICU admission. Firstly, a promising score from a large-scale study in China appears to perform poorly when applied to a European cohort, mainly by lack of specificity. Secondly, biological features that are quite significant for the admission to ICU such as LDH (cut-off 579 UI/l) or venous lactate (cut-off 3,02 mmol/l) cannot predict death, suggesting that these two outcomes should be clearly distinguished. Nevertheless, in our cohort we report a significant NLR cut-off of 22.1. Thirdly, the percentage of respiratory impairment (which is not significant for admission to ICU, cf. Table 6) is significant to predict death.

## Conclusions

A predictive score for admission to ICU or death is urgently needed in secondary hospitals such as Clinique Saint-Pierre Ottignies. Interleukine-6 (IL-6) has been shown as a marker of severity of the disease. Meta-analysis of mean IL-6 concentrations demonstrated three-fold higher levels in patients with complicated COVID-19 compared with patients with noncomplicated disease^27^. However, most developed countries hospitals cannot afford costly laboratory exams. Optimal allocation of resources guided by clinical-based indicators will best guide patients at time of admission and avoid futile treatments in intensive care units. These indicators are still lacking. Wynants *et al*.^5^ have shown that proposed models in the literature are at high risk of bias and their reported performances probably optimistic. We show that a predictive score based on a large scale population study^1^ may not be reliable enough to be implemented in Belgium, both for admission to the ICU and for death, mainly because of lack of specificity. However, in our small cohort it appears that LDH above 579 UI/l and venous lactate above 3.02 mmol/l may be considered as good predictive biological factors for ICU admission. On the other side, death risk can be assessed by NLR above 22.1, tobacco abuse status and <u>a 80% of lung injury</u>. The development of reliable predictive methods are of great importance to ensure fair allocation of scarce medical resources in time of COVID-19^28^. However, our results suggest that available methods may not have to require accuracy to be beneficial in emergency ward.

## Data Availability

All data relevant to the study are included. No further data available.

